# Nusinersen in adult patients with 5q spinal muscular atrophy: a multicenter observational cohorts’ study

**DOI:** 10.1101/2021.06.15.21258262

**Authors:** Juan F Vázquez-Costa, Mónica Povedano, Andrés E Nascimiento-Osorio, Antonio Moreno Escribano, Solange Kapetanovic Garcia, Raul Dominguez, Jessica M Exposito, Laura González, Carla Marco, Julita Medina Castillo, Nuria Muelas, Daniel Natera de Benito, Nancy Carolina Ñungo Garzón, Inmaculada Pitarch Castellano, Teresa Sevilla, David Hervás

## Abstract

**Objective:** To assess safety and efficacy of nusinersen in adult 5q spinal muscular atrophy (SMA) patients.

**Methods:** Patients older than 15 years and followed at least for 6 months with one motor scale (Hammersmith Functional Motor Scale Expanded, HFMSE; Revised Upper Limb module, RULM) in five referral centers were included. Clinical and patients’ global impression of change (CGI-C and PGI-C) were recorded in treated patients at the last visit. Functional scales (Egen Klassification, EK2; Revised Amyotrophic Lateral Sclerosis Functional Rating Scale, ALSFRS-R) and the percent-predicted forced vital capacity were collected when available.

**Results:** Seventy-nine SMA patients (39 treated with nusinersen) were included. Compared with untreated patients, treated patients showed a significant improvement of 2 points (±0.46) in RULM (p<0.001) after six months. After a mean follow-up of 16 months, nusinersen treatment was associated with a significant improvement in HFMSE (OR=1.15, p=0.006), 6MWT (OR=1.07, p<0.001), and EK2 (OR=0.81, p=0.001).

Compared with untreated patients, more treated patients experienced clinically meaningful improvements in all scales, but these differences were statistically significant only for RULM (p=0.033), ALSFRS-R (p=0.005), and EK2 (p<0.001). According to the CGI-C and PGI-C, 64.1% and 61.5% of treated patients improved with treatment. Being non-sitter was associated with less response to treatment, while longer time of treatment was associated with better response. Most treated patients (77%) presented at least one adverse event, mostly mild.

**Conclusions:** Nusinersen treatment associates to some improvements in adult SMA patients. Most severely affected patients with complex spines are probably those with the most unfavorable risk-benefit ratio.

## Introduction

5q spinal muscular atrophy (SMA) is a genetic neurodegenerative disease, caused by a homozygous deletion or mutation in the survival motor neuron1 (*SMN1*) gene, affecting the lower motor neurons (LMN). This results in progressive tetraparesis, affecting first to lower limbs and later to upper limbs, followed by respiratory insufficiency, dysarthria and dysphagia.^1,2^ According to the age of symptoms onset and to the highest acquired motor milestone, SMA children are typically classified in type 1-3. SMA type 1 patients will never be able to sit unsupported, while SMA type 2 patients will never be able to walk independently.^3^ SMA types, and therefore the disease severity, are largely explained by the number of *SMN2* gene copies, which is also capable to produce a small amount of SMN protein.^4^ Thus, while SMA type 1 patients will usually die during the childhood, most type 2 and 3 patients will reach the adulthood with a variable degree of disability.^5^ The rare type 4 patients typically start after 30 years old and will not present any noteworthy disability.^1^ Due to the disease progression, the SMA type, defined in the infancy, does not reliably inform about the functionality in the adulthood. Therefore, adult SMA patients are functionally classified in non-sitters, sitters and walkers.^3^

Nusinersen, an antisense oligonucleotide, was approved for the treatment of SMA after being shown to improve survival and motor function in infants and children in two randomized placebo-controlled clinical trials.^6,7^ Conversely, in the adolescent and adult population, the evidence of nusinersen efficacy is based on real world studies, usually small case series, with controversial results.^8–12^ Moreover, functional and patient’s reported outcome (PRO) data on nusinersen efficacy have been scarcely reported,^13,14^ despite its importance for regulatory agencies. Considering the high frequency of adverse events (AEs) associated with repeated lumbar punctures and the high costs of the treatment, it is of utmost importance to add real world evidence of nusinersen efficacy in the adult population. Therefore, the objective of this study was to report the safety as well as motor and functional outcomes in a multicentre Spanish cohort of treated and non-treated adult SMA patients.

## Methods

### Study design and participants

Nusinersen was approved in Spain for the treatment of SMA patients in March 2018, with some restrictions posed by a protocol of the Spanish health department.^15^ Briefly, very severe (defined as EK2 > 47, or requiring non-invasive ventilation –NIV-for more than 16 hours a day) or mild (type 3 patients with HFSME > 54, or type 4) SMA patients were usually excluded from treatment.

For this prospective observational study, SMA patients from 5 centers in Spain were included (Hospital la Fe, Hospital Sant Joan de Deu, Hospital de Bellvitge, Hospital Virgen de la Arrixaca, Hospital de Basurto). Inclusion criteria were: a) genetically confirmed SMA (either homozygous deletion or compound heterozygous mutation in *SMN1*); b) older than 15 years at the baseline visit; c) longitudinal data on at least one motor scale at the time of the study closure (August 2020). Patients meeting the criteria established by the health department, were routinely offered nusinersen treatment. The final decision to start the treatment was made by the patient after discussion with the neurologist of pros and cons. Since the protocol approval, prospective data of treated and untreated patients were collected at baseline, 6 months later and every 6-12 months later on. When available, retrospective data of untreated patients were also collected from October 2015.

### Procedures

Treated patients were injected with the 12 mg loading doses of nusinersen (at days 0, 14, 28 and 65) and maintenance doses every 4 months, as per label. Conventional and imaging-guided (including ultrasound,^16^ fluoroscopy and CT) lumbar punctures were performed by experienced neurologists and neuroradiologists, respectively. All treated patients received at least four doses of nusinersen, except one patient,^16^ who was discontinued after the second dose of nusinersen due to the lack of lumbar access and was excluded from efficacy analysis.

Motor and functional scales were administered by experienced and/or trained neurologists and physiotherapists. All centers collected the same motor scales and pulmonary tests, but functional scales were missing in some centers. Moreover, not all scales are applicable to all patients (see below). Consequently, the number and characteristics of SMA patients varies in each scale.

### Clinical variables and outcomes

Age, gender, and age at symptom’s onset, as well as the presence of severe scoliosis (>45º Cobb angle) and/or scoliosis surgery were recorded in all the patients upon recruitment. Patients were classified in type 1 to 4 as defined elsewhere,^1^ as well as in functional subgroups:^3^ walkers (able to walk at least 5 steps without assistance), sitters (able to sit without assistance nor head support for more than 10 seconds) and non-sitters. The use of NIV, gastrostomy and salbutamol was also recorded at baseline in all patients.

The following outcome measures were used to assess efficacy.

The Hammersmith Functional Motor Scale Expanded (HFMSE) consists in 33 items, with a maximum of 66 points (higher scores indicating better function), and it is designed for the assessment of sitters and walkers.^17^ Based on natural history data and patients interviews, a score change of more than 2 points is considered to be clinically meaningful.^17,18^

The Revised Upper Limb Module (RULM), includes 20 items with a maximum score of 37 (higher scores indicating better function).^19^ It has been validated in both ambulant and non-ambulant patients, and a score change of 2 points or more is considered to be clinically meaningful.^8,12^

The 6-minutes walk test (6MWT) measures the distance a patient is able to walk within 6 minutes, and it is therefore only applicable to walkers. Based on previous clinical trial data in Duchenne patients, a change of 30 meters or more was considered to be clinically meaningful.^20^

The Egen Klassification 2 (EK2) is a functional scale that includes 17 items on 8 daily-life categories (wheelchair use, wheelchair transfers, trunk mobility, eating, swallowing, breathing, coughing, fatigue). Each item is scored from 0 to 3 for a maximum of 51 points (higher scores indicating worse function). It has been designed for and validated in non-ambulant SMA population.^21,22^

The Revised Amyotrophic Lateral Sclerosis Functional Rating Scale (ALSFRS-R) is a functional scale that includes 12 items on 4 domains (bulbar, upper limbs, lower limbs, respiratory). Each item is scored from 0 to 4 for a maximum of 48 points (higher scores indicating better function). It was designed for ALS patients, but it has also been used in adult SMA patients,^9,23^ in whom it has been recently validated (manuscript sent for publication).

According to their specific validity, the 6MWT was assessed in walkers, the HFMSE in walkers and sitters, and the EK2 in sitters and non-sitters. The RULM, ALSFRS-R and the percent-predicted forced vital capacity (FVC%) were assessed in all subgroups of patients.

Furthermore, the clinical and the patients’ global impression of change (CGI-C and PGI-C) was obtained in all treated patients at the last visit. For the CGI-C, neurologists were asked to respond to the following question about each patient: “ compared to his/her condition right before treatment, how much has the patient changed?” For the PGI-C, patients were asked to respond to the following question: “ compared to your condition before treatment, how are you doing overall?” Responses were collected in a semi-quantitative manner from very much worse (−3), to very much improved (+3), with 0 being no change.

To assess safety, following items were recorded systematically in each visit: the patient-reported adverse events (AEs), categorized by severity and relationship to treatment; the start of NIV or placement of gastrostomy; abnormal routine laboratory findings.

### Statistical analysis

Data were summarized as means, standard deviations, medians, and first and third quartiles for the continuous variables, and as relative and absolute frequencies for the categorical variables.

Rank-based regression models were used to analyze the effect of treatment on the visit scores at 6 months. For these models, the baseline scores and the treatment with nusinersen were included as predictive variables. To analyze the effect of treatment on the visit scores at the last visit, mixed ordinal regression models were used. Since last visit comprises different time intervals in each patient and the effect of treatment is expected to increase with time,^8,12^ both the follow up time (in months) and the interaction between time and treatment were included as predictive variables.

Convergence problems appeared in the fitted ordinal regression models of two scales (ALSFRS-R and RULM), due to our limited sample size. Bayesian modelling adjustment with a weakly informative prior (N(0, 3)) were used in those cases. For each model, only the estimate of the effect of treatment is shown (Table 2 and 3).

For the calculation of the responders’ rate, several definitions of responder were used. Firstly, the percentage of treated and untreated patients that improved at least the minimal clinically important difference (MCID) established for each scale was calculated. For the EK2 and ALSFRS-R scales a change ≥ 2 points were considered as clinically meaningful, based on the investigators’ experience. Chi square tests were used to assess the differences in responder rates as defined above. Secondly, we measured the percentage of treated patients who experienced at least mild improvements (1 point) according to the CGI-C and the PGI-C.

We also assessed the concordance between CGI-C and PGI-C using the Bangdiwala’s observer agreement chart for ordinal variables.^24^ A weight of 1 was settled up for a complete agreement and a weight of 0.5 for a partial agreement, defined as a difference of 1 point between CGI-C and PGI-C. Differences between scores > 1 points were considered as disagreement. The agreement was quantified as moderate when B = 0.50 to 0.69, strong when rs = 0.70 to 0.89 and very strong when rs = 0.90 to 1.00.

Finally, an ordinal multivariable model was used to assess those variables predicting improvement according to the CGI-C.

All analyses were pre-specified before looking at the data. P values < 0.05 were considered statistically significant. All the statistical analyses and graphs were performed with the R software (version 4.0.3).

### Ethical approval

The study was approved by the Ethics Committee for Biomedical Research of Instituto de Investigación Sanitaria la Fe and Fundació Sant Joan de Déu. All the participants gave written informed consent.

### Data availability

All data supporting our findings are available on reasonable request.

## Results

### Population characteristics

The study included 79 SMA patients (39 treated with nusinersen). Their demographic and clinical characteristics are summarized in Table 1. Treated patients were somewhat older (33 vs 30 years old), and more frequently male (51% vs 42%) and type 3 (74% vs 42%). Untreated patients were more frequently non-sitter (50% vs 26%) and NIV users (38% vs 23%), despite shorter disease duration (25 vs 29 years). Both subgroups had a similar rate of concomitant salbutamol treatment.

**Table 1.**
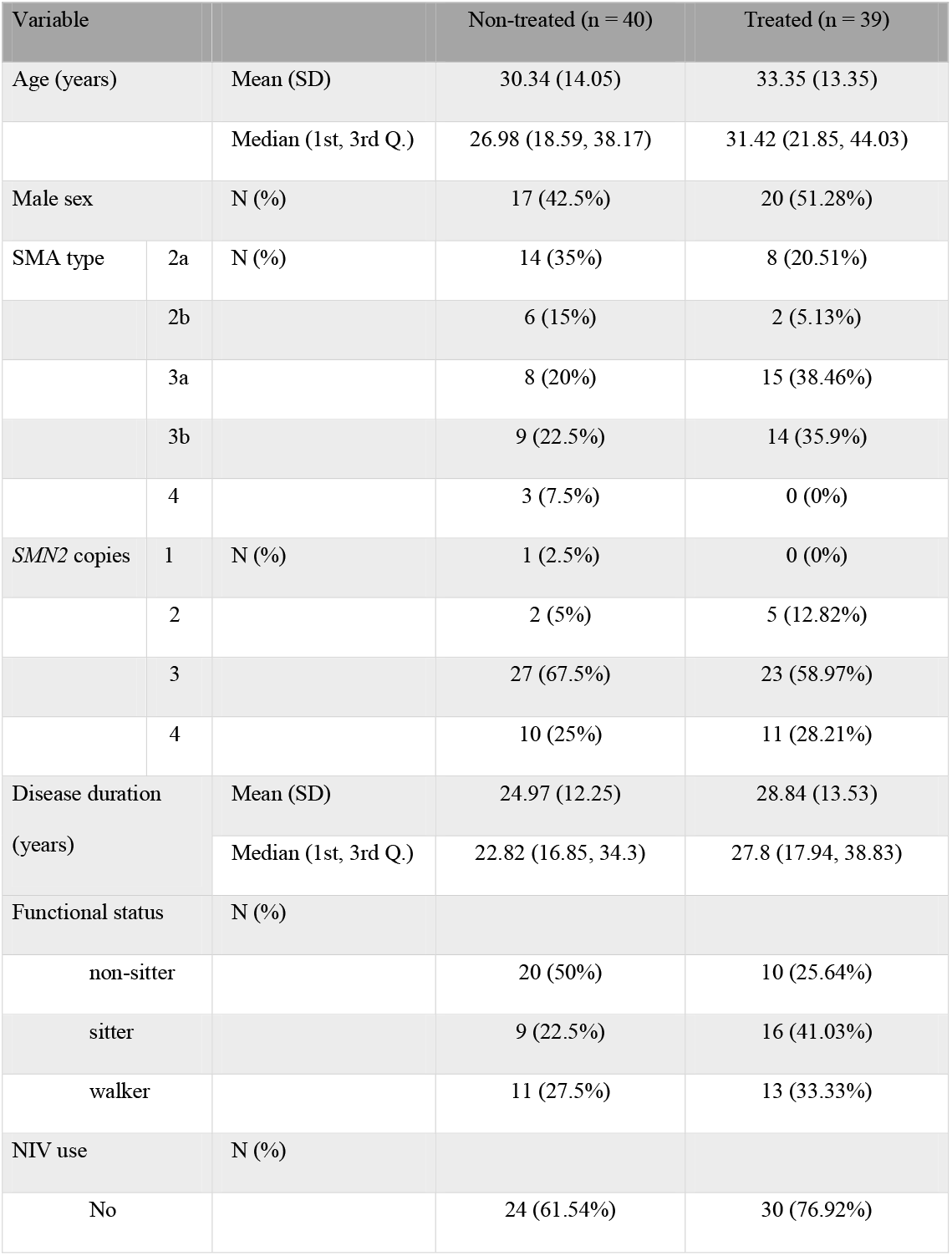
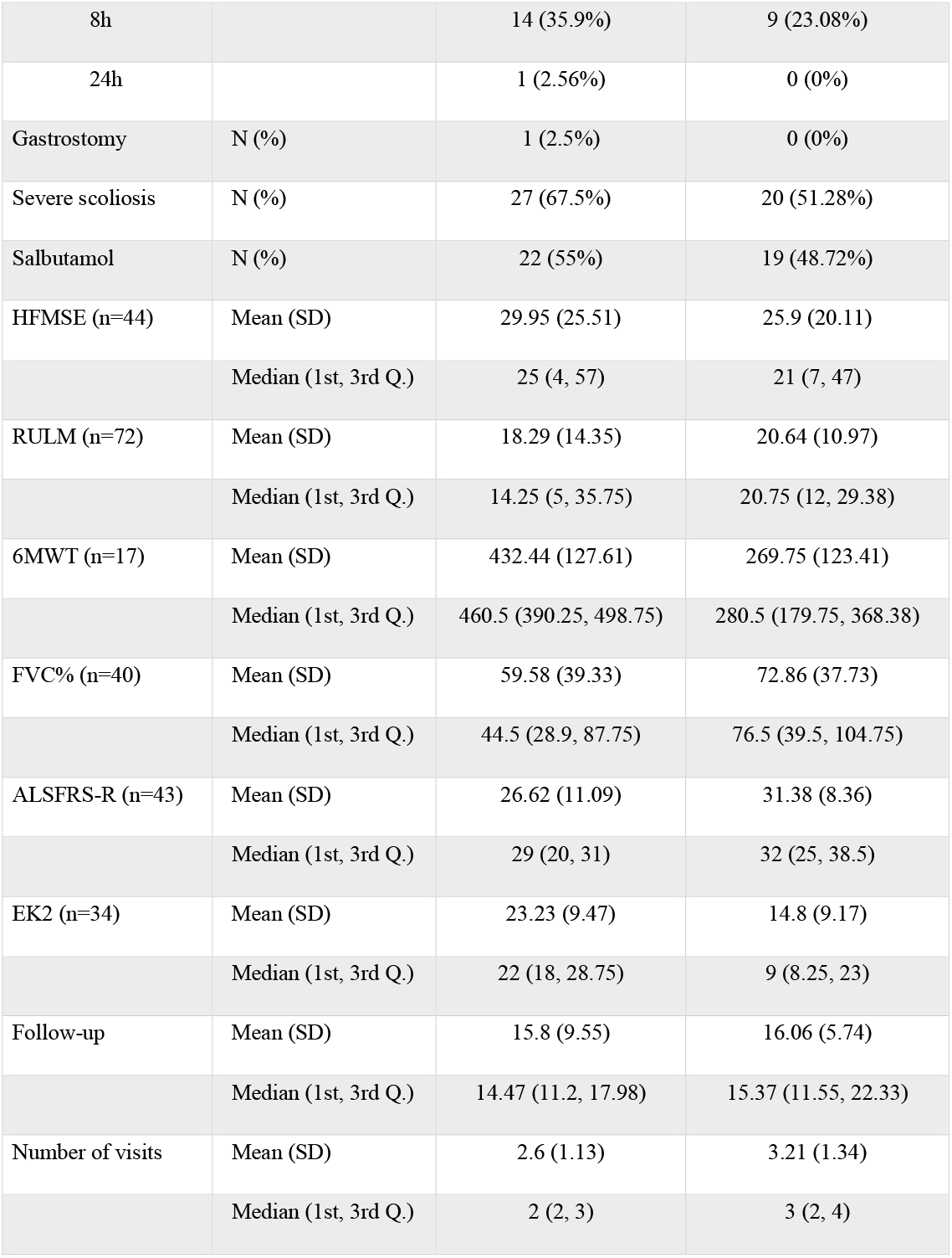
Demographical and baseline clinical characteristics of SMA patients included in the study. ALSFRS-R: Revised version of the Amyotrophic Lateral Sclerosis Functional Scale; EK2: Egen Klassifikation 2; FVC%: Percent-predicted Forced Vital Capacity; HFMSE: Hammersmith Functional Motor Scale Expanded; RULM: Revised Upper Limb Module; 6MWT: 6-Minutes Walk Test.

| Variable |  |  | Non-treated (n = 40) | Treated (n = 39) |
| --- | --- | --- | --- | --- |
| Age (years) |  | Mean (SD) | 30.34 (14.05) | 33.35 (13.35) |
|  |  | Median (1st, 3rd Q.) | 26.98 (18.59, 38.17) | 31.42 (21.85, 44.03) |
| Male sex |  | N (%) | 17 (42.5%) | 20 (51.28%) |
| SMA type | 2a | N (%) | 14 (35%) | 8 (20.51%) |
|  | 2b |  | 6 (15%) | 2 (5.13%) |
|  | 3a |  | 8 (20%) | 15 (38.46%) |
|  | 3b |  | 9 (22.5%) | 14 (35.9%) |
|  | 4 |  | 3 (7.5%) | 0 (0%) |
| SMN2 copies | 1 | N (%) | 1 (2.5%) | 0 (0%) |
|  | 2 |  | 2 (5%) | 5 (12.82%) |
|  | 3 |  | 27 (67.5%) | 23 (58.97%) |
|  | 4 |  | 10 (25%) | 11 (28.21%) |
| Disease duration<br>(years) |  | Mean (SD) | 24.97 (12.25) | 28.84 (13.53) |
|  |  | Median (1st, 3rd Q.) | 22.82 (16.85, 34.3) | 27.8 (17.94, 38.83) |
| Functional status |  | N (%) |  |  |
| non-sitter |  |  | 20 (50%) | 10 (25.64%) |
| sitter |  |  | 9 (22.5%) | 16 (41.03%) |
| walker |  |  | 11 (27.5%) | 13 (33.33%) |
| NIV use |  | N (%) |  |  |
| No |  |  | 24 (61.54%) | 30 (76.92%) |

Table 1. Demographical and baseline clinical characteristics of SMA patients included in the study.
|  |  |  |  |
| --- | --- | --- | --- |
| 8h |  | 14 (35.9%) | 9 (23.08%) |
| 24h |  | 1 (2.56%) | 0 (0%) |
| Gastrostomy | N (%) | 1 (2.5%) | 0 (0%) |
| Severe scoliosis | N (%) | 27 (67.5%) | 20 (51.28%) |
| Salbutamol | N (%) | 22 (55%) | 19 (48.72%) |
| HFMSE (n=44) | Mean (SD) | 29.95 (25.51) | 25.9 (20.11) |
|  | Median (1st, 3rd Q.) | 25 (4, 57) | 21 (7, 47) |
| RULM (n=72) | Mean (SD) | 18.29 (14.35) | 20.64 (10.97) |
|  | Median (1st, 3rd Q.) | 14.25 (5, 35.75) | 20.75 (12, 29.38) |
| 6MWT (n=17) | Mean (SD) | 432.44 (127.61) | 269.75 (123.41) |
|  | Median (1st, 3rd Q.) | 460.5 (390.25, 498.75) | 280.5 (179.75, 368.38) |
| FVC% (n=40) | Mean (SD) | 59.58 (39.33) | 72.86 (37.73) |
|  | Median (1st, 3rd Q.) | 44.5 (28.9, 87.75) | 76.5 (39.5, 104.75) |
| ALSFRS-R (n=43) | Mean (SD) | 26.62 (11.09) | 31.38 (8.36) |
|  | Median (1st, 3rd Q.) | 29 (20, 31) | 32 (25, 38.5) |
| EK2 (n=34) | Mean (SD) | 23.23 (9.47) | 14.8 (9.17) |
|  | Median (1st, 3rd Q.) | 22 (18, 28.75) | 9 (8.25, 23) |
| Follow-up | Mean (SD) | 15.8 (9.55) | 16.06 (5.74) |
|  | Median (1st, 3rd Q.) | 14.47 (11.2, 17.98) | 15.37 (11.55, 22.33) |
| Number of visits | Mean (SD) | 2.6 (1.13) | 3.21 (1.34) |
|  | Median (1st, 3rd Q.) | 2 (2, 3) | 3 (2, 4) |

Overall, better baseline scores were found in treated vs untreated patients (Table 1) except in the 6MWT (because none of the type 4 patients were treated) and in the HFMSE (because it was not assessed in non-sitters).

Treated patients received a mean of 6 doses of nusinersen and 45% of them required imaging-guided lumbar puncture.

### Treatment effect at 6 months

At 6 months, an improvement in treated patients was predominant in all scales and tests, while in untreated patients, scores usually worsened or remained stable except for the 6MWT (Figure 1). Nusinersen treatment, adjusting by baseline scores, improved 2 points (±0.46) in RULM (p<0.001) according to the model, but differences in other scales were not statistically significant (Table 2).

**Figure 1.**
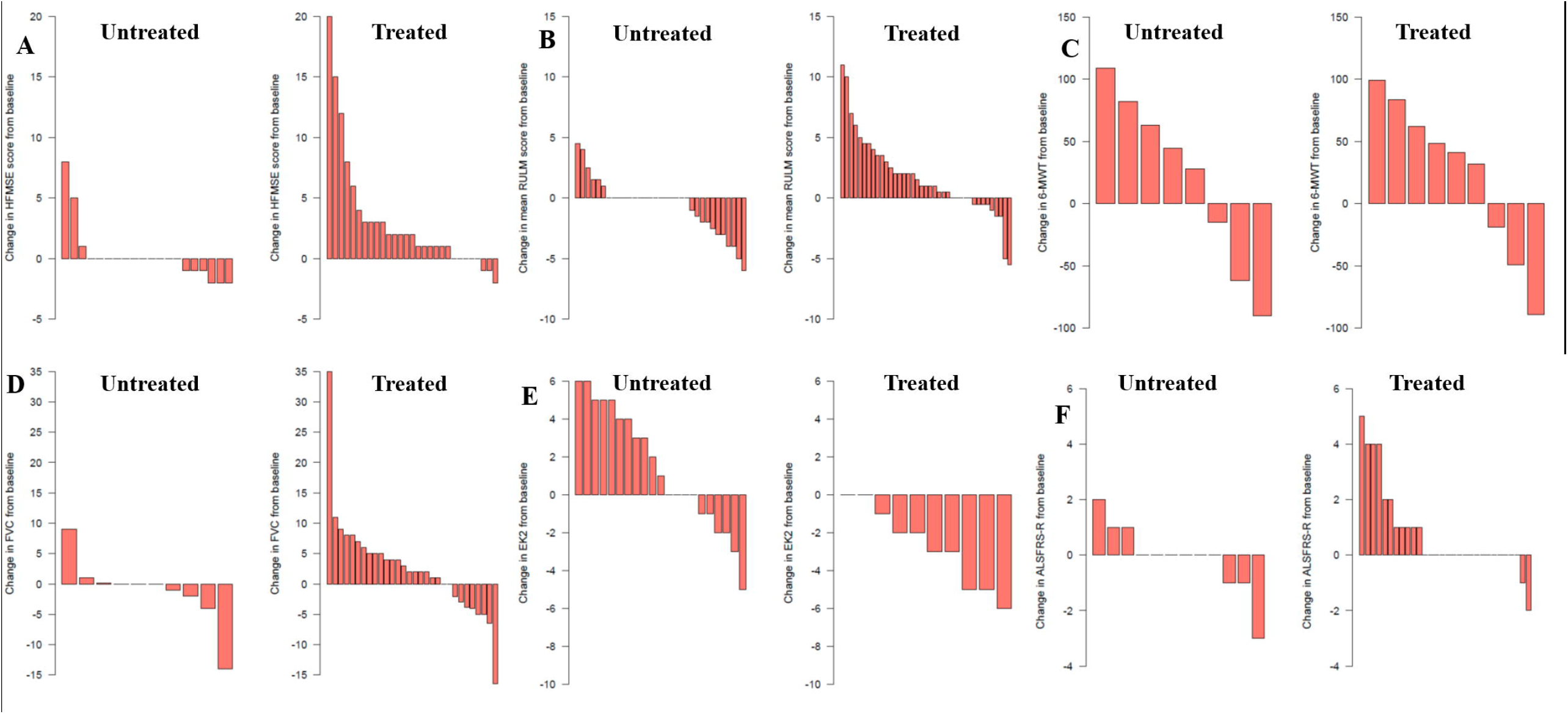
Individual changes in scores from baseline to T1 (6 months) in treated and untreated patients in the different tests: A) HFMSE; B) RULM; C) 6MWT; D) FVC%; E) EK2; F) ALSFRS-R. ALSFRS-R: Revised version of the Amyotrophic Lateral Sclerosis Functional Scale; EK2: Egen Klassifikation 2; FVC%: Percent-predicted Forced Vital Capacity; HFMSE: Hammersmith Functional Motor Scale Expanded; RULM: Revised Upper Limb Module; 6MWT: 6-Minutes Walk Test.

**Table 2.** Raw score differences between baseline and 6 months’ visits in treated and untreated patients and the estimated effect of nusinersen according to the multivariable model, after adjusting for baseline values. In bold, statistically significant results. ALSFRS-R: Revised version of the Amyotrophic Lateral Sclerosis Functional Scale; EK2: Egen Klassifikation 2; FVC%: Percent-predicted Forced Vital Capacity; HFMSE: Hammersmith Functional Motor Scale Expanded; RULM: Revised Upper Limb Module; 6MWT: 6-Minutes Walk Test. *p<0.001

| Test | Raw scores |  | Estimate | SE | p |
| --- | --- | --- | --- | --- | --- |
|  | Untreated | Treated |  |  |  |
| HFMSE (n=44) | 0.16 (1.83) | 2.43 (4.52) | 2 | 1.12 | 0.082 |
| **RULM (n=71)** | **-0.58 (2.27)** | **1.67 (3.28)** | **2** | **0.46** | **<0.001\*** |
| 6MWT (n=17) | 19.94 (70.03) | 23.22 (62.75) | -6.27 | 46.12 | 0.894 |
| FVC% (n=40) | -1.09 (5.65) | 2.6 (8.29) | 3.19 | 2.11 | 0.139 |
| ALSFRS-R (n=42) | -0.08 (1.24) | 0.77 (1.59) | 3.42 | 3.03 | 0.999 |
| EK2 (n=30) | 1.07 (2.83) | -2.72 (2.74) | -4 | 3.19 | 0.221 |

### Treatment effect at the last visit

Both, treated and untreated patients were followed up for a mean of 16 months (Table 1), albeit more visits were performed in treated patients (3.21 vs 2.6). At the last visit, after adjusting for the baseline values and follow-up time, the effect of treatment was associated with a significant improvement in HFMSE (OR=1.15 IC 95% [1.04, 1.27], p=0.006), 6MWT (OR=1.07 IC 95% [1.06, 1.08], p<0.001), and EK2 (OR=0.81 IC 95% [0.71, 0.92], p=0.001) and a non-statistically significant improvement was found in all other scales (Table 3).

**Table 3.** Effect of the interaction “treatment and follow-up time” in the different outcomes at the last visit available for each scale. In bold, statistically significant results. P values are lacking in variables calculated with Bayesian models. ALSFRS-R: Revised version of the Amyotrophic Lateral Sclerosis Functional Scale; EK2: Egen Klassifikation 2; FVC%: Percent-predicted Forced Vital Capacity; HFMSE: Hammersmith Functional Motor Scale Expanded; RULM: Revised Upper Limb Module; 6MWT: 6-Minutes Walk Test. *p<0.001

|  | OR | Lower 95 | Upper 95 | p |
| --- | --- | --- | --- | --- |
| **HFMSE (n=44)** | **1.15** | **1.041** | **1.271** | **0.006** |
| RULM (n=71) | 1.022 | 0.961 | 1.091 | - |
| **6MWT (n=17)** | **1.071** | **1.065** | **1.078** | **<0.001\*** |
| FVC% (n=40) | 1.002 | 0.9 | 1.116 | 0.975 |
| ALSFRS-R (n=42) | 1.036 | 0.94 | 1.144 | - |
| **EK2 (n=31)** | **0.809** | **0.712** | **0.92** | **0.001** |

### Responders and variables predicting response

According to the MCID of each scale a variable percentage of treated patients (25% - 80%) experienced clinically meaningful improvements at the last visit (Table 4). However, some patients not treated with nusinersen also experienced clinically meaningful improvements (Table 4). Compared with untreated patients, more treated patients experienced clinically meaningful improvements in all scales, but these differences were statistically significant only for RULM, ALSFRS-R and EK2 (Table 4).

**Table 4.**
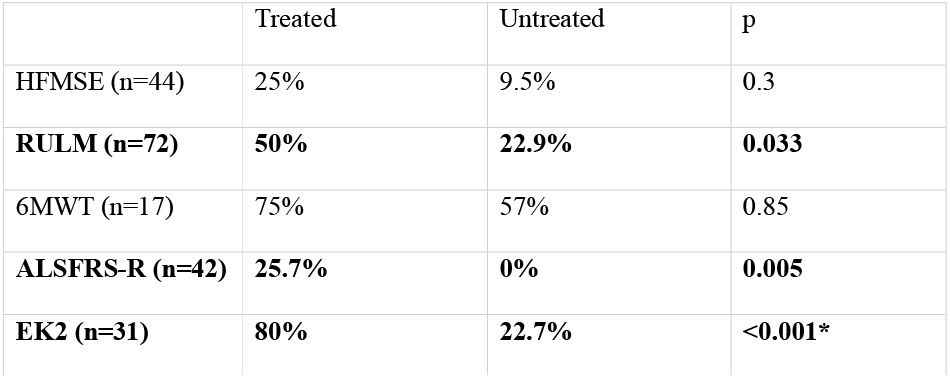
Percentage of patients experiencing clinically meaningful impairments (as defined in methods) in each scale at the last visit. ALSFRS-R: Revised version of the Amyotrophic Lateral Sclerosis Functional Scale; EK2: Egen Klassifikation 2; HFMSE: Hammersmith Functional Motor Scale Expanded; RULM: Revised Upper Limb Module; 6MWT: 6 Minutes Walk Test. *p<0.001

|  | Treated | Untreated | p |
| --- | --- | --- | --- |
| HFMSE (n=44) | 25% | 9.5% | 0.3 |
| **RULM (n=72)** | **50%** | **22.9%** | **0.033** |
| 6MWT (n=17) | 75% | 57% | 0.85 |
| **ALSFRS-R (n=42)** | **25.7%** | **0%** | **0.005** |
| **EK2 (n=31)** | **80%** | **22.7%** | **<0.001\*** |

According to the CGI-C and PGI-C, 64.1% and 61.5% of treated patients improved, while 0% and 2.5% of patients respectively deteriorated (Figure 2). There was a high agreement between CGI-C and PGI-C (unweighted agreement 0.6, weighted agreement 0.8).^24^ A CGI-C of 3 (very much improved) was scored in two SMA type 3a patients. A sitter with 4 *SMN2* copies, who was able to stand still with help but had lost her ability to walk some years before, improved 20 points in HFMSE, 10 points in RULM and was able to walk unaided 30 meters in the 6MWT after 14 months of treatment. Another walker with 3 *SMN2* copies who had been deteriorating the year before treatment start and was close to lose ambulation, improved 24 points in HFMSE, 7 points in RULM and 183 meters in 6MWT, after 14 months of treatment (supplementary figure 1).

**Figure 2.**
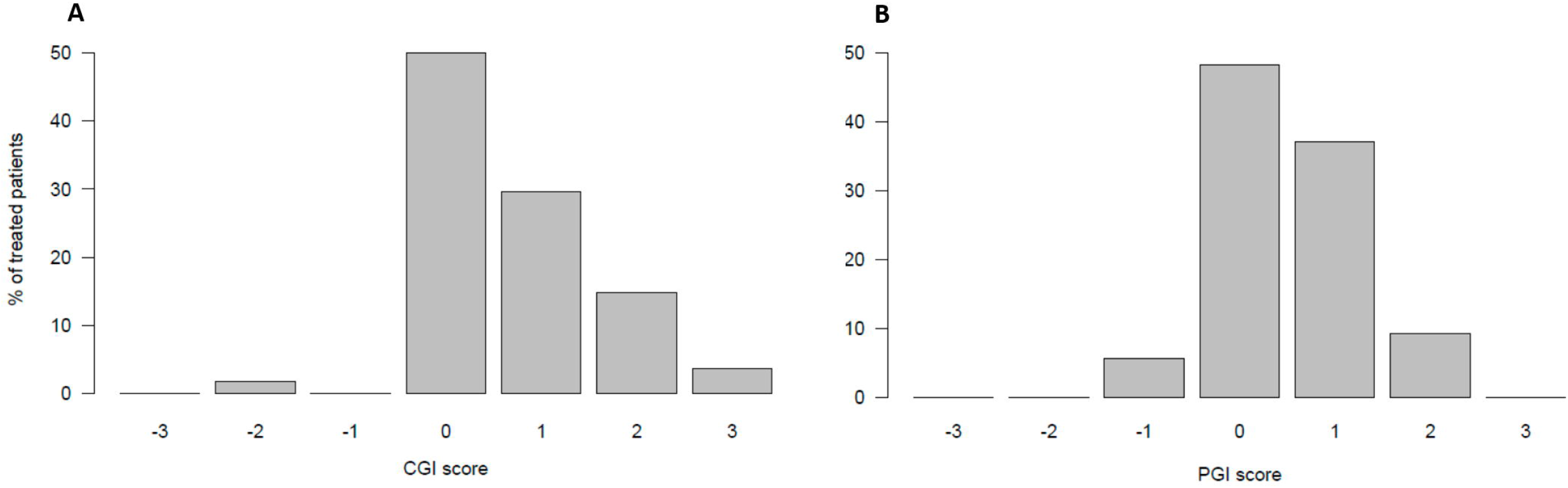
Graphical representation of the clinical global impression of change (CGI-C) and the patient’s global impression (PGI-C) scores.

According to the multivariable model (Table 5), being non-sitter (compared with walker) was associated with less response to treatment, as assessed with the CGI-C, while longer time of treatment was associated to better response.

**Table 5.** Multivariable model assessing the effect of several variables in the response to treatment, as defined per the clinical global impression of change scale. In bold, statistically significant results.

|  | Estimate | SE | P |
| --- | --- | --- | --- |
| Age | - 0.043 | 0.035 | 0.226 |
| Disease duration | 0.047 | 0.034 | 0.175 |
| *SMN2* copy number | 0.183 | 0.369 | 0.623 |
| Sitter | -0.382 | 0.36 | 0.297 |
| **Non-sitter** | **-0.912** | **0.406** | **0.032** |
| **Treatment duration** | **0.054** | **0.024** | **0.035** |

### Adverse events

Thirty treated patients (77%) presented at least one AE during the follow-up. Overall, 55 AEs were reported, mostly related with the administration procedure: 45 were mild (post-lumbar puncture syndrome and lumbar pain) and 10 were moderate (7 post-lumbar puncture syndrome, 2 urinary retention due to neurogenic bladder, 1 radial neurapraxia). Two patients (5%) discontinued treatment due to adverse events (repeated post-lumbar puncture syndromes) and another due to technically challenging lumbar punctures. One treated patient started NIV during follow-up, after a respiratory infection that required hospitalization. No clinically relevant laboratory changes were found.

## Discussion

This multicenter study provides class III evidence that nusinersen improves motor function in at least a subset of SMA patients, and causes frequent, usually mild, adverse events.

Since the approval of nusinersen for the treatment of SMA patients, it has been widely used in the adult population, despite the absence of clinical trials assessing its efficacy and safety in this subgroup of patients. Moreover, previous research has largely overlooked the particularities of adult SMA patients, characterized by a huge clinical heterogeneity and poorly defined natural history.^25,26^

Recently, the first reports of real-world evidence on the efficacy and safety of nusinersen in adult SMA patients have been published.^8–14^ Most of them, but not all,^10^ suggest nusinersen efficacy, at least in a subset of patients. However, they show several pitfalls that difficult their interpretation.^27^ Firstly, many of them are small case series. Secondly, treated patients were usually followed up for less than two years and a direct comparison with a control group of untreated patients was lacking. While any improvement in a neurodegenerative disease could be regarded as a treatment effect, previous natural history studies have shown that individual improvements in some motor scales in a timeframe less of that two years are not infrequent in the adult population.^18,28–31^ Moreover, in the last years, treatments such as salbutamol or pyridostigmine are frequently used off-label for the treatment of SMA patients and could have a positive effect in motor scales,^32^ erroneously attributed to nusinersen.

Accordingly, a recent systematic review highlighted that the scarcity of data, the phenotypic variability, slow disease progression and the limited sensitivity of available outcome measures difficult to draw definitive conclusions about the natural history of the disease in short timeframes.^26^ Thirdly, in previous studies, patients were frequently stratified following the classical children classification instead of as functional subgroups, as previously recommended.^3,26,33^ Moreover, HFMSE was a common outcome for all patients in those studies, despite not being designed to assess non-sitter patients.^33^ Finally, functional scales and PROs have been scarcely used to describe treatments effects, despite their importance in the clinical practice and for regulatory agencies.

This multicenter study used real-world data to assess nusinersen efficacy and safety, while overcoming some previous methodological limitations. Namely, a control group with natural history data was included for direct comparison and, importantly, a similar percentage of patients were treated with salbutamol in both the control and the nusinersen group. Moreover, patients were categorized in functional subgroups, in which validated motor and functional scales as well as PROs were appropriately used. Finally, the statistical approach was designed to control for common pitfalls in real world studies, such as selection bias and the variability in the follow-up.

Overall, our results support previous evidence suggesting the efficacy of nusinersen. Thus, after 6 months of treatment, treated patients showed an improvement of 2 points in RULM compared with untreated patients, and this difference was statistically significant after adjusting by baseline scores. Moreover, improvements were found in other motor and functional scales although, due to the heterogeneity of the sample and the limited sample size, differences were not statistically significant.

This positive effect was confirmed at the last visit, after a mean follow-up of 16 months, when nusinersen treatment was independently associated with statistically significant improvements in HFMSE, 6MWT and EK2 and a positive, but non-significant effect, in RULM and ALSFRS-R. Overall, the effect of treatment in motor scales, as showed in our models, seem to be modest, in line with previous studies.^8,9,12^ Interestingly, the greatest effect was found in EK2, a bedside functional scale for the assessment of non-ambulant patients. This could reflect its ability to detect mild functional changes in non-ambulant patients and to measure the effect of nusinersen on fatigability, which has been previously reported after nusinersen treatment^34,35^ and might not be captured by HFMSE and RULM. However, direct comparisons between scales should be interpreted with caution for two reasons: the small sample size and the variable use of scales among the sample, according to the functional subgroup and the center.

Remarkably, those outcome measures applicable to all functional subgroups (RULM, ALSFRS-R and FVC%) failed to show statistically significant improvements. This suggests that the measurement of treatment effect in real-world studies is also hindered by the huge heterogeneity of SMA patients. Thus, whenever possible, functional stratification should be considered in studies addressing adult SMA patients.^25,33^ Previous studies have reported a 30-60% of responders, according to the predefined MCID of motor scales.^8,11,12^ However, the responder rate should also be interpreted with caution, since two important biases could lead to under- and overestimations.

On the one hand, both HFMSE and RULM show floor and ceiling effects,^12,28,36^ which could reduce their sensitivity to detect changes in more mildly and severely affected patients. The use of functional scales showing higher sensitivity to changes, such as EK2 or ALSFRS-R, could increase the responder rate. Thus, in our study, the rate of responders ranged from 25% of treated patients according to the HFMSE and 80% of treated patients according to EK2.

On the other hand, our and previous natural history studies show that a non-negligible proportion of “ untreated” adult patients experience improvements that could be considered clinically meaningful, when followed for less than two years.^18,28–31^ These unexpected improvements could be due to three facts: test-retest inaccuracies in the scales; functional fluctuations, which are frequently reported by patients (e.g. depending on the season of the year); or to the fact that some “ untreated” patients have actually started other treatments (e.g. salbutamol, physical therapy, etc…) during or right before the study. Thus, the comparison with a control group can help to interpret the results. In our study, the responder rate was greater in all scales in treated vs untreated patients, but this difference was statistically significant only for RULM and EK2.

Finally, PROs such as CGI-C and PGI-C have been widely used to assess the responder rates in both clinical trials and real-world studies. According to them, about 60-65% of treated patients experienced at least minimal clinically meaningful improvements. This includes about 25% of patients experiencing moderate improvements, with two of them showing remarkable improvements of over 20 points in HFMSE. Interestingly, SMA children also showed a variable response to nusinersen in clinical trials.^37–39^ Younger age (which in children is closely related to shorter disease duration), better baseline functionality, and more *SMN2* copies were associated with better response to treatment in those trials.^37–39^ In adults, better baseline functionality has been the only factor suggested to correlate with greater improvement in motor scales.^8,12^ However, given the floor effect of motor scales, it could be argued that the improvements experienced by patients with minimal functionality are not adequately captured. Our multivariable model, based on the GIC (which captures both objective and subjective improvements), confirmed that non-sitters are less probable to respond to treatment, while age, disease duration and the *SMN2* copy number did not seem to influence the response. Moreover, longer treatment duration was associated with greater response, in keeping with previous reports.^8,12^

It has been claimed that the mild improvements found in the adult SMA population after nusinersen treatment could be due to placebo effect.^10^ While placebo effect might indeed explain some improvements, increasing evidence supports also a physiological effect of nusinersen. Firstly, unlike it would be expected in a placebo effect, the improvement of patients increased with time of treatment.^8,12^ Secondly, some patients experienced huge improvements that are neither spontaneously expected nor explicable in a neurodegenerative disease. Finally, large series (including this) show pretty consistent results in functional and motor scales, and PROs.^8,11,12^

Nevertheless, when deciding to start a treatment, the potential benefit must be balanced against the risks and the costs of treatment. We and others have shown that nusinersen treatment in adult SMA patients is associated with a high frequency (30-80%) of AEs.^8,10–12,14^ While most are mild and transient, some of them are permanent (neurogenic bladder, radiation exposure), can be life-threatening (meningitis, subarachnoid hemorrhage),^10,40^ or lead to short-term treatment discontinuation (7.7% in our series). Most AEs are related to the administration procedure and could be more frequent and severe in patients with complex spines,^1040^ in whom transforaminal approaches are frequently tried. The use of non-traumatic needles and ultrasound-guided parasagittal approaches^16^ could help to reduce the frequency and severity of AEs. Moreover, the use of reservoirs and port devices for the intrathecal administration of nusinersen could improve the risk-benefit ratio but should be first evaluated in the setting of clinical trials.

While the decision to start any treatment should be made at an individual level, our and previous studies suggest that most disabled patients (i.e., non-sitters) are less likely to improve with nusinersen, being also probably those with greater risks of serious AEs. Therefore, the use of nusinersen in these patients should be evaluated carefully, especially considering the availability of oral alternatives.

If non-improving patients treated with nusinersen will benefit from long-term stabilization or not, especially in terms of respiratory impairment and survival, should be clarified in future studies. Moreover, given their greater body mass index, adult SMA patients could potentially respond better to higher nusinersen doses. This hypothesis will be tested in a small subgroup of adult patients in the DEVOTE study (NCT04089566).

Notwithstanding the strengths of our study, it also has several limitations, which are common in real-world studies in rare diseases. A greater sample size would have been desirable to be able to stratify the results according to the functional subgroups and to increase the power of the multivariable analysis. Hence, our study might be underpowered to detect some positive effects. Moreover, despite a common protocol, there was some methodological heterogeneity among centers, especially regarding retrospective data. Thus, not all patients were visited at the same intervals, and functional scales and FVC were not routinely administered in all patients. Furthermore, baseline patients’ characteristics were somewhat different in treated and untreated groups, since most severe and mild patients were not treated as per protocol. However, the statistical analysis was designed to minimize all these limitations, for example by adjusting by baseline scores and the follow-up time.

In conclusion, our multicenter real-world study provides class III evidence that nusinersen treatment associates with mild motor and functional improvements in up to 60% of adult SMA patients, but also causes frequent mild adverse events. Most severely affected patients with complex spines are probably those with the most unfavorable risk-benefit ratio. Collaborative real-world studies are warranted to improve the prediction of which patients will benefit from each treatment and why. This becomes increasingly important considering the huge cost of new treatments and the low class of evidence available for adult SMA patients.

## Data Availability

All data supporting our findings are available on reasonable request.

## Data availability

JFVC and DH had full access to the database population used to create the study population. All data supporting our findings are available on reasonable request.

## Acknowledgments

We would like to thank patients and patients’ associations (FundAME, GaliciAME and ForzAME) for their collaboration in this study. We also thank Fernando Mora, M Carmen Baviera, Sandra Roca and Obdulia Moya for their participation in patients’ assessment.

---

| **Name** | **Location** | **Contribution** |
| --- | --- | --- |
| Juan F Vázquez-Costa | Hospital Universitario y Politécnico la Fe, Valencia | Designed the study, participated in clinical data acquisition and interpretation, and wrote and edited the manuscript |
| Mónica Povedano | Bellvitge Hospital-IDIBELL | Clinical data acquisition and interpretation, and critically revised the manuscript |
| Andrés Nascimento-Osorio | Hospital Sant Joan de Déu, Barcelona | Clinical data acquisition and interpretation, and critically revised the manuscript |
| Antonio Moreno Escribano | Hospital Clínico Universitario Virgen de la Arrixaca, Murcia, Spain | Clinical data acquisition and interpretation, and critically revised the manuscript |
| Solange Kapetanovic Garcia | Hospital Universitario Basurto - OSI Bilbao, Spain | Clinical data acquisition and interpretation, and critically revised the manuscript |
| Raul Dominguez | Bellvitge Hospital-IDIBELL | Clinical data acquisition |
| Jessica M Exposito | Hospital Sant Joan de<br>Déu, Barcelona | Clinical data acquisition |
| Laura González | Bellvitge Hospital-IDIBELL | Clinical data acquisition |
| Carla Marco | Bellvitge Hospital-IDIBELL | Clinical data acquisition |
| Julita Medina<br>Castillo | Hospital Sant Joan de<br>Déu, Barcelona | Clinical data acquisition and<br>interpretation, and critically revised the<br>manuscript |
| Nuria Muelas | Hospital Universitario<br>y Politécnico la Fe,<br>Valencia | Critically revised the manuscript |
| Daniel Natera | Hospital Sant Joan<br>de Déu, Barcelona | Clinical data acquisition and<br>interpretation, and critically revised the<br>manuscript |
| Nancy Carolina<br>Ñungo Garzón | Hospital Universitario<br>y Politécnico la Fe,<br>Valencia | Clinical data acquisition and<br>interpretation, and critically revised the<br>manuscript |
| Inmaculada<br>Pitarch-Castellano | Hospital Universitario<br>y Politécnico la Fe,<br>Valencia | Clinical data acquisition and<br>interpretation, and critically revised the<br>manuscript |
| Teresa Sevilla | Hospital Universitario<br>y Politécnico la Fe,<br>Valencia | Critically revised the manuscript |
| David Hervás | Universitat<br>Politècnica de<br>València, Valencia | Planned and performed statistical analysis<br>and interpretation, and critically revised<br>the manuscript |

Supplementary figure 1. Graphical representation of the scores of HFMSE, RULM and 6MWT in a walker SMA patient before and after nusinersen treatment. HFMSE: Hammersmith Functional Motor Scale Expanded; RULM: Revised Upper Limb Module; 6MWT: 6 Minute Walk Test.

