## Supplementary figures and images for "Nusinersen in adult patients with 5q spinal muscular atrophy: a multicenter observational cohorts’ study"

### Suplementary figure 1

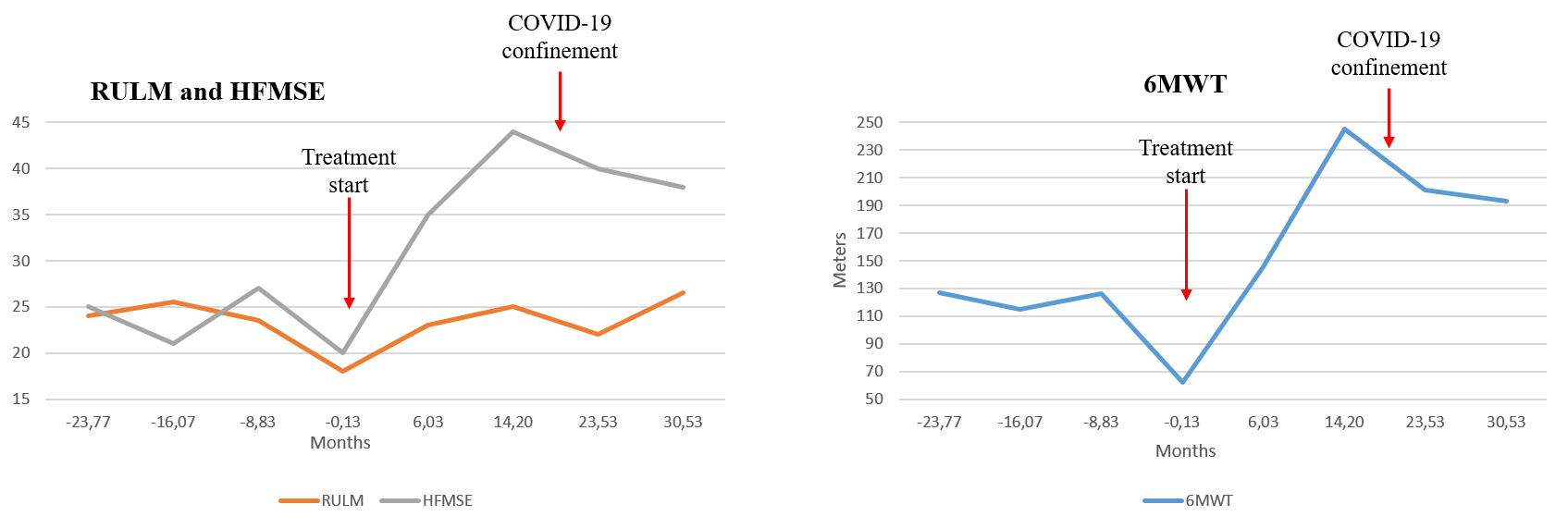
